# Hepatitis B core-related antigen (HBcrAg) point-of-care tests as a risk stratification tool: experience from Kenya

**DOI:** 10.1101/2024.11.25.24316890

**Authors:** Louise O Downs, Dorcas Okanda, Oscar Chirro, Mwanakombo Zaharani, Benson Safari, Nadia Aliyan, Monique I Andersson, Yasuhito Tanaka, Anthony O Etyang, Yusuke Shimakawa, George Githinji, Philippa C Matthews

## Abstract

We undertook a point-of-care test (POCT) for hepatitis B core-related antigen (HBcrAg) on adults living with HBV in Kilifi, Kenya. A positive test identified all with hepatitis B viral load >200,000 IU/ml, who were HBeAg positive, and correlated with higher ALT (p = 0.03), raised APRI (p<0.001) and elastography scores (p = 0.03).

## Introduction

The 2024 Global Hepatitis Report highlights significant morbidity and mortality due to chronic hepatitis B virus infection (CHB), with the number of people dying from CHB rising from 820,000 in 2019 to 1.1 million in 2022, disproportionately affecting populations in the WHO African Region (WHO-AFRO) (1).

One of the most significant barriers to HBV elimination in this region is that only 0.2% of people living with HBV (PLWHB) are on treatment (1) and tests for risk-stratification are often inaccessible and unaffordable. New 2024 WHO HBV management guidelines simplify assessment, but still recommend alanine aminotransferase (ALT) measurement or aspartate transaminase (AST) to platelet ratio index (APRI) calculation (2) along with HBV viral load (VL) measurement to determine long-term treatment eligibility. HBV VL measurement and/or hepatitis B e antigen (HBeAg) assessment are also recommended to determine eligibility for peri-partum antiviral prophylaxis (3).

Point-of-care tests (POCTs) offfer an attractive alternative to these laboratory assays, allowing real-time feedback, potentially improving retention in clinical care and viral suppression rates (4). Currently, liver stiffness measurement using elastography is the only validated POCT for assessing liver health for PLWHB; however, the hardware is prohibitively expensive. Current POCTs for HBeAg have poor sensitivity (5,6), and HBV VL measurements require a laboratory platform like Gene Xpert (Cepheid Inc., CA, USA).

Hepatitis B core-related antigen (HBcrAg) correlates with peripheral HBV DNA and HBeAg status in untreated individuals (7) and could be an alternative to HBV VL quantification in low resource settings (8). HBcrAg has previously only been available as a chemiluminescent assay (CLEIA). A new HBcrAg POCT performed well in The Gambia as a surrogate for HBV VL, but has not yet been assessed elsewhere in the general adult population with CHB in WHO-AFRO countries, and performance has not been evaluated against ALT or liver fibrosis. We set out to evaluate the HBcrAg-POCT in PLWHB in Kilifi, Kenya, considering (i) the ease of testing, (ii) its relationship with ALT, HBeAg status, HBV VL and elastography, and (iii) its contribution to determining treatment eligibility.

## Methods

PLWHB were recruited through the STRIKE-HBV study at KEMRI-Wellcome Trust Research Programme (KWTRP), which identified 102 non-pregnant adults from Kilifi County, Kenya as previously described (10). Positive HBsAg POCT results were confirmed with an enzyme linked immunoassay (ELISA) (Murex HBsAg 3, Diasorin). The cohort was 60% female, median age 37 years (IQR 28-49 years), and two people were also living with HIV infection.

Liver elastography was undertaken at recruitment (Echosens, Paris, France), and blood samples were drawn and transported to the research laboratory within 2-3 hours of collection, spun to separate serum, and frozen in aliquots at −80°C. Routine laboratory markers were measured in a validated diagnostic clinical laboratory at KWTRP, including ALT, AST (Ilab Aries) and platelets (AcT 5diff CP, Beckman Coulter) for APRI score calculation.

HBeAg was tested at the KWTRP research laboratory using CLEIA (Ig Biotechnology, CA, US), HBV VL was tested at Oxford University Hospitals, UK using the Abbott Alinity (Illinois, USA). The HBcrAg-POCT (ESPLINE™ (RUO), Fujirebio, Japan) was shipped from Kumamoto University, Japan, stored at room temperature and undertaken retrospectively on 50µls defrosted serum in the KWTRP laboratories as per manufacturer’s instructions (9); no specific training was required. The testing was performed in three batches by clinical and laboratory staff, blinded to patient details including HBV VL and HBeAg status. Positive and negative controls were provided and undertaken once prior to each batch testing.

Liver health thresholds and treatment eligibility criteria were taken from the 2024 WHO HBV guidelines (2) as follows: elastography score >7.0kPa = significant fibrosis, >12.5kPa = cirrhosis; APRI >0.5 = significant fibrosis, >1.0 = cirrhosis. Those with i) elastography score >7kPa; OR ii) APRI score >0.5; OR iii) HBV DNA >2000 IU/ml AND ALT > upper limit of normal (ULN, 19 IU/L for women, 30 IU/L for men), were defined as treatment eligible. In the untreated population, we first investigated whether ALT alone could identify those who were treatment eligible, then evaluated the contribution of the HBcrAg POCT.

Statistical analysis was done using R version 4.2.0. The study was approved by the Scientific Ethics Review Unit (SERU 4565) and Oxford Tropical Network Ethics Committee (OxTREC 23-22).

## Results

HBcrAg-POCT was positive in 14/102 PLWHB (14%) – with a positivity rate of 4/61 (7%) in women and 10/41 (24%) in men ((p=0.01). Those with a positive-POCT were younger (median age 28 years compared to 38 years with a negative-POCT, p=0.03). There was no difference in current antiviral treatment status between those testing POCT-positive or negative (table 1). Laboratory personnel reported the POCT twas easy to perform. The test turnaround time was approximately 40 minutes.

**Table 1:** Characteristics of 102 adults with chronic HBV infection in Kilifi, Kenya, assessed with a point of care test (POCT) for Hepatitis B core-related antigen (HBcrAg).

| Characteristic (all participants) | N=102 | Negative HBcrAg POCT<br>n = 88<sup>1</sup> | Positive HBcrAg POCT<br>n = 14<sup>1</sup> | p-value<sup>2</sup> |
| --- | --- | --- | --- | --- |
| **Sex** | 102 |  |  | **0.010** |
| Female | 61 | 57 (93%) | 4 (7%) |  |
| Male | 41 | 31 (76%) | 10 (24%) |  |
| **Age in years, median (IQR)** | 102 | 38 (30, 49) | 28 (27, 39) | **0.03** |
| Unknown | 1 |  |  |  |
| **On Antiviral Therapy** | 102 |  |  | 0.8 |
| Yes | 27 | 24 (89%) | 3 (11%) |  |
| No | 75 | 64 (85%) | 11 (15%) |  |
| Characteristic (untreated group) | N=75 | Negative HBcrAg POCT<br>n = 64<sup>1</sup> | Positive HBcrAg POCT<br>n = 11<sup>1</sup> | p-value<sup>2</sup> |
| **HBV DNA (log<sub>10</sub> IU/ml), median (IQR)** | 74 | 2.68 (1.56, 3.45) | 6.05 (3.19, 7.03) | **<0.001** |
| **HBV DNA Group (log<sub>10</sub> IU/ml)** | 75 |  |  | **<0.001** |
| <20 | 13 | 12 (92%) | 1 (8%) |  |
| 20 - 2000 | 36 | 34 (94%) | 2 (6%) |  |
| 2000 - 20,000 | 15 | 14 (93%) | 1 (7%) |  |
| 20,000 - 200,000 | 3 | 3 (100%) | 0 (0%) |  |
| >200,000 | 7 | 0 (0%) | 7 (100%) |  |
| Unknown | 1 | 1 | 0 |  |
| **ALT U/L, median (IQR)** | 75 | 26 (19, 35) | 58 (31, 67) | **0.006** |
| **ALT >ULN** | 75 |  |  | 0.7 |
| Yes | 48 | 40 (83%) | 8 (17%) |  |
| No | 27 | 24 (89%) | 3 (11%) |  |
| **Elastography score (kPa), median (IQR)** | 66 | 4.25 (3.50, 5.55) | 6.65 (3.50, 8.98) | 0.2 |
| Unknown | 9 | 6 | 3 |  |
| **Liver Health Fibroscan** | 75 |  |  | **0.03** |
| Normal ($\leq 7$ kPa) | 57 | 52 (91%) | 5 (9%) | |
| Significant fibrosis ($> 7 - \leq 12.5$ kPa) | 8 | 6 (75%) | 2 (25%) | |
| Cirrhosis ($> 12.5$ kPa) | 1 | 0 (0%) | 1 (100%) | |
| Unknown | 9 | 6 | 3 |  |
| **APRI Score, median (IQR)** | 74 | 0.29 (0.22, 0.39) | 0.57 (0.34, 0.86) | **0.003** |
| Unknown | 1 | 1 | 0 |  |
| **Liver health APRI** | 75 |  |  | **<0.001** |
| Normal ($< 0.5$) | 60 | 57 (95%) | 3 (5%) | |
| Significant Fibrosis (0.5 - 1) | 11 | 5 (45%) | 6 (55%) |  |
| Cirrhosis ($> 1$) | 3 | 1 (33%) | 2 (67%) | |
| Unknown | 1 | 1 | 0 |  |
| **HBeAg** | 75 |  |  | **<0.001** |
| Positive | 7 | 0 (0%) | 7 (100%) |  |
| Negative | 68 | 64 (94%) | 4 (6%) |  |

<sup>1</sup>n (%); Median (IQR), <sup>2</sup>Pearson's Chi-squared test; Wilcoxon rank sum test; Fisher's exact test
IQR - interquartile range, ALT - alanine aminotransferase, ULN - upper limit of normal, kPa - kilopascals,
APRI - AST to platelet ratio index, HBeAg - hepatitis B 'e' antigen.

### Analysis in the untreated population

75 PLWHB were untreated on cohort entry (42 female, 33 male). Median age was 40 years (IQR 27 - 49) among whom 11/75 (15%) had a positive HBcrAg POCT. This group had a significantly higher median HBV DNA level than those with a negative POCT (6.05 log_10_IU/ml vs 2.7 log_10_IU/ml, respectively; p <0.001, table 1). To identify those testing HBV DNA >200,000 IU/ml, HBcrAg-POCT had a sensitivity of 100% (7/7) (supp figure 1) and a specificity of 94% (64/68). All those who had HBV DNA >200,000 IU/ml also carried HBeAg (table 1).

A positive HBcrAg-POCT was significantly associated with a higher median ALT than a negative test (58 IU/L vs 26 IU/L, p=0.006, table 1). Individuals with a positive HBcrAg-POCT were more likely to meet elastography criteria for significant fibrosis or cirrhosis than those with a negative POCT (supp figure 1, 3/11, (27%) vs 6/64 (9%) with a negative test, p=0.03), but there was no significant difference in median elastography score between those with a positive and negative HBcrAg-POCT. Those with a positive HBcrAg-POCT had higher median APRI scores (0.57 vs 0.29, p = 0.003), and were more likely to meet APRI criteria for significant fibrosis or cirrhosis (supp figure 1, 8/11(73%) vs 6/63, (10%), respectively; p<0.001 (table 1).

In this currently untreated group, 27/75 (36%) people met treatment criteria as defined. Among these, 23/27 (85%) would have been identified based on ALT criteria alone, whereas a positive HBcrAg-POCT alone would have only identified 9/27 (33%) (supp table 1). Of the four people not identified by ALT, one had a positive HBcrAg-POCT, but the other three did not, so would have been missed by both tests. Therefore the addition of HBcrAg POCT to ALT identified one extra treatment eligible person compared to using ALT alone (supp. figure 2). This individual also had a high HBV VL of 6.1 log_10_IU/ml and fibroscan score of 10.1kPa, suggesting some existing liver fibrosis and a risk of progressive chronic liver disease.

## Discussion

This is the first report directly comparing the HBcrAg-POCT with ALT, markers of liver fibrosis, and HBeAg status. Here, a positive HBcrAg-POCT was strongly associated with liver inflammation by ALT, and fibrosis/cirrhosis by both Fibroscan and APRI scores. However, the HBcrAg-POCT alone identified fewer people meeting treatment eligibility scores by APRI or Fibroscan than ALT alone. Addition of the POCT to ALT did identify one extra person as treatment eligible who was at high risk of complications, however further assessment is needed to see if this benefit translates to larger populations.

In this cohort, a positive HBcrAg-POCT was 100% sensitive at identifying HBeAg positivity, performing much better than currently available HBeAg-POCTs which have sensitivities ranging from 30-62% (5,6). A positive HBcrAg-POCT was 100% sensitive at identifying those with HBV VL >200,000 IU/ml, which is a higher sensitivity than what was found when this POCT was trialled in the Gambia. This POCT is therefore of potential clinical utility to identify individuals eligible for perinatal antiviral prophylaxis. This is the first time this has been shown in East Africa, where circulating HBV genotypes differ from those in West Africa.

This HBcrAg-POCT is quick and could be undertaken without specific training. It is relatively low production cost (<$5 per test), compared to HBV VL which varies in price across WHO AFRO up to $62/test (1), and HBeAg up to $40/test for the laboratory ELISA (11,12) assuming these tests are actually available. ALT is relatively low cost (<$10) and available in much of WHO-AFRO, so continues to be a good method of liver health assessment in the absence of other tests. Further implementation research is needed to determine whether inclusion of the HBcrAg-POCT alongside ALT could enhance linkage to care and improve decentralisation of clinical assessment, and how its impact and cost-effectiveness varies by setting.

The number of PLWHB included in this study was small, from a specific geographical region and did not include children/adolescents or pregnant women. We did not have access to a HBeAg POCT or gold standard method of assessing liver disease such as cross-sectional imaging or liver biopsy.

## Conclusion

HBcrAg-POCT correlates strongly with several liver disease markers in PLWHB. However, it does not perform better than ALT at identifying those with abnormal elastography or APRI scores, so should be used as an adjunct rather than stand-alone. It could provide a low-cost alternative to some otherwise unavailable, unaffordable diagnostics.

## Acknowledgements

We thank Fujirebio Inc. for providing test reagents and to all the staff at the Comprehensive Care Clinic, outpatients department and Maternal Child Health Clinics at Kilifi County Referral Hospital for assisting in study recruitment.

## Author Contributions

LOD, DO, YS and PCM conceptualised the manuscript

LOD, PCM, NA, MIA, AOE and GG designed the study

LOD, OC, MZ, BS and NA enrolled patients and collected study data and serum samples

DO and LOD undertook the point of care tests in the KWTRP laboratories

YS and YT provided the point of care test kits and gave technical advice and oversight on the use of the point of care tests.

LOD undertook data analysis with advice provided by AOE, YS, GG and PCM.

All authors reviewed and commented on the manuscript prior to submission.

## Data Availability

For the purpose of Open Access, the author has applied a CC-BY public copyright license to any author accepted manuscript version arising from this submission. Data supporting the findings of this study will be publicly available on the acceptance of the manuscript for publication. This manuscript was written with the permission of Director KEMRI CGMRC.

---

**Supplementary Table 1:** The number of people living with hepatitis B virus infection in Kilifi, Kenya meeting treatment criteria (defined as Fibroscan score >7kPa, or APRI score >0.5, or the combination of raised ALT AND HBV DNA >2000 IU/ml) compared with how many of these people would have been identified using ALT alone, HBcrAg-POCT alone or by both tests. APRI: Aspartate transaminase-to-platelet ratio index, ALT: Alanine aminotransferase, HBcrAg-POCT: Hepatitis B core-related antigen point-of-care test. Treatment criteria: Fibroscan score >7kPa or APRI score >0.5.

| Meeting treatment criteria | Identified by ALT alone | Identified by HBcrAg-POCT alone | Identified by ALT AND HBcrAg-POCT together |
| --- | --- | --- | --- |
| 27/75 (36%) | 23/27 (85%) | 9/27 (33%) | 24/27 (89%) |

**Supplementary figure 1:**
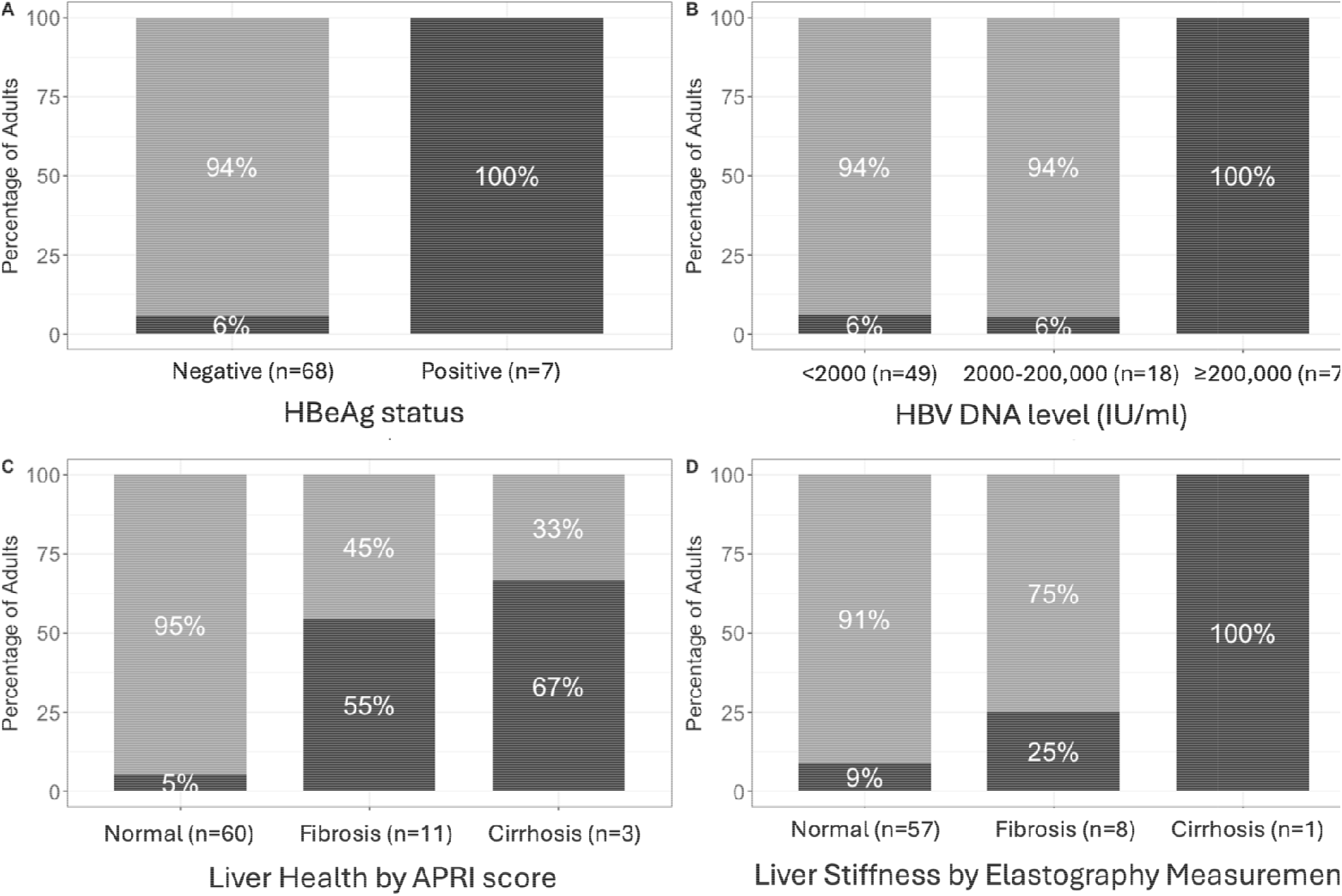
Relationship between clinical characteristics and results of HBcrAg point-of-care test in adults living with hepatitis B virus infection (HBV) in Kilifi, Kenya: A - HBeAg status, B - HBV VL level, C - liver health based on APRI scores, D - Liver stiffness by elastography scores. Each characteristic is split by hepatitis B core-related antigen (HBcrAg) POCT test result. Light grey is those testing HBcrAg POCT negative, dark grey is those testing HBcrAg POCT positive. HBeAg: hepatitis B ‘e’ antigen, VL: viral load, APRI: Aspartate transaminase to platelet ratio index. APRI scores: Normal <0.5, Significant fibrosis 0.5 - 1, Cirrhosis >1. Elastography scores: Normal <7 kPa, Significant fibrosis 7-12.5 kPa, Cirrhosis >12.5 kPa.

**Supplementary figure 2:**
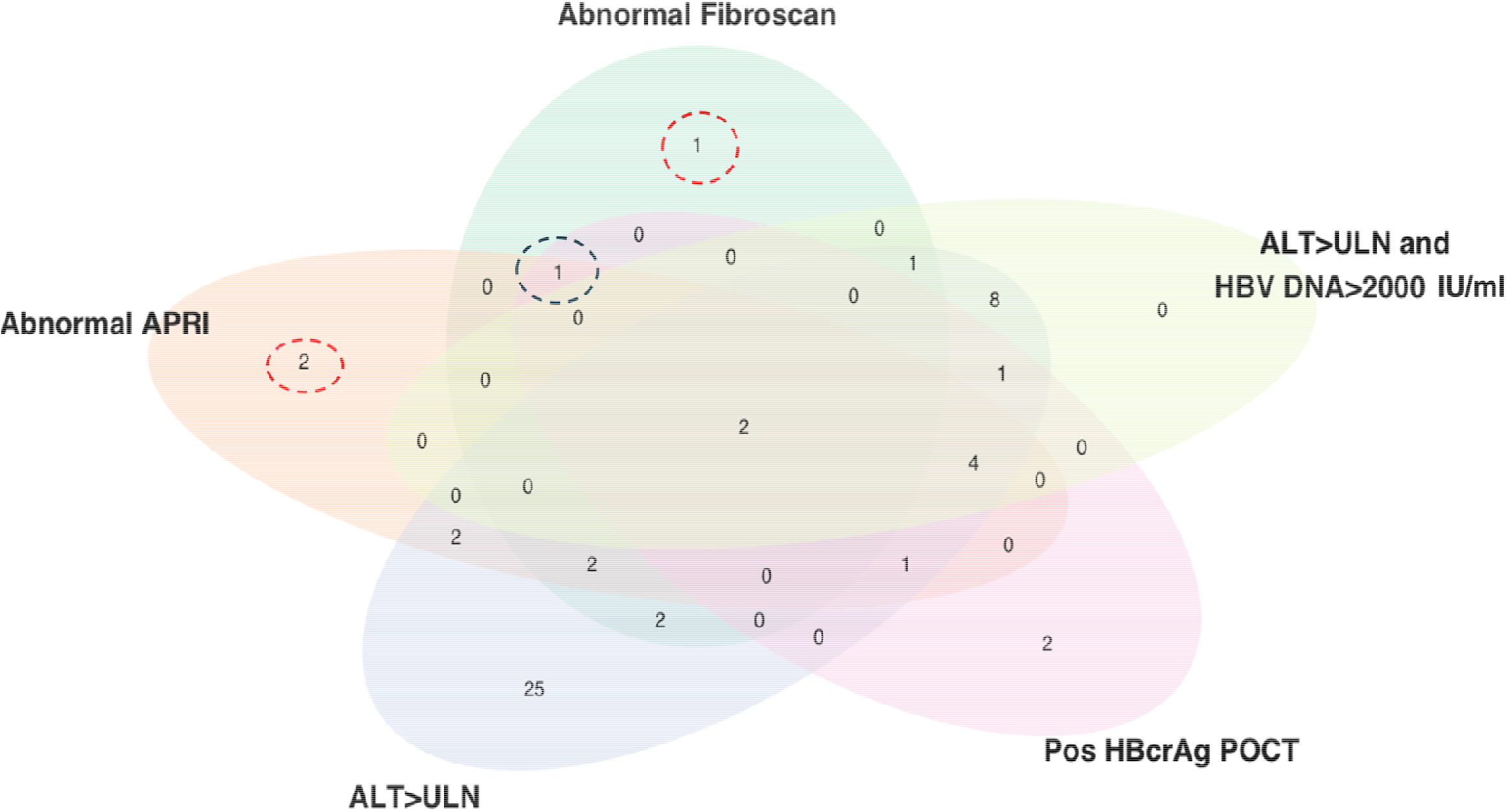
Venn diagram showing numbers of untreated adults living with hepatitis B infection who would be eligible for nucleoside analogue therapy based on different criteria. Black dotted circle indicates the one extra person with abnormal APRI and elastography scores who would have been identified using HBcrAg-POCT but missed using abnormal ALT alone. Red dotted circles indicate the three people with abnormal Fibroscan or APRI scores that would have been missed by both ALT and HBcrAg-POCT.

